# The impact of behavioral interventions on respiratory viruses in a two-cohort study

**DOI:** 10.1101/2024.05.06.24306945

**Authors:** Behnam Nikparvar, Cara Exten, Nita Bharti

## Abstract

Behavioral interventions are a critical tool for managing novel and emerging pathogens. However, the dynamics of behavioral interventions have been difficult to measure and are poorly understood. In this study, we investigate the uptake, persistence, and waning, of behavioral interventions among two cohorts in Centre County, PA, focusing on the three years following the emergence of SARS-CoV-2. We detected clusters of behaviors that followed similar patterns of engagement and variations over time and identified some novel COVID-19 behavioral interventions that may have severely disrupted non-COVID-19 respiratory disease incidence. Additionally, we detected links between changes in risk perception and changes in behaviors over time. These findings can inform recommendations around behavioral interventions during outbreak management, including information dissemination and behavioral guidelines.

## 1. Introduction

Behavioral interventions (BIs), also known as non-pharmaceutical interventions (NPIs), can play a critical role in managing infectious diseases. They are especially important when managing novel or emerging pathogens (Gamma et al., 2019; Manguvo & Mafuvadze, 2015; Markel et al., 2007), for which pharmaceutical resources are limited or nonexistent (Knock et al., 2021; Perra, 2021). Extensive research conducted during the COVID-19 pandemic estimated that BIs likely reduced SARS-CoV-2 transmission significantly (Flaxman et al., 2020; Lai et al., 2020) as well as the incidence of other respiratory viruses, including seasonal influenza (Huh et al., 2021; Kraay et al., 2021). Although the impact of BIs on respiratory diseases is extensively supported by literature, their uptake, persistence, and waning are not well understood.

BIs are diverse and their uptake and persistence are impacted by many factors. Examples of BIs include practicing personal hygiene, maintaining physical distance, wearing facemasks, following quarantine and isolation recommendations, regulating the sizes of gatherings, working remotely, closing schools, and restricting travel. In the United States, some BIs, such as maintaining physical distance, wearing facemasks, or regulating the sizes of the gatherings were newly implemented in 2020 following the emergence of SARS-CoV-2 while others, such as practicing personal hygiene and, to some degree, staying home when feeling ill, have been consistently promoted (Smith et al., 2023). During the COVID-19 pandemic, the targeted behaviors for BI strategies changed to adapt to an evolving situation resulting in distinct dynamics for each behavior.

BIs are often layered to amplify their impact (Hatchett et al., 2007, Lai et al., 2020) and recent empirical research shows that BIs may be adopted as interconnected packages rather than isolated actions (Smith et al., 2023). This raises the question of whether this tendency to cluster is only in the adoption or to a larger extent in dynamics (adoption, persistence, and waning) of BIs. BI engagement depends on cognitive factors (e.g., risk perception), psychosocial factors (e.g., anxiety, social support, fatigue from practicing BIs), socioeconomics (e.g., income and education), access to information, and demographics (e.g., age and gender) (Raude et al., 2020, Endalew et al., 2022; Leung et al., 2022). Among these factors, risk perception or perceived susceptibility is frequently the focal point of health behavior change interventions. Recent findings, including research on the COVID-19 pandemic, indicate that an increase in perceived risk leads to engagement in behaviors that reduce the risk of infection (Brewer et al., 2004; Bruine de Bruin & Bennett, 2020; Dryhurst et al., 2022; Ferrer & Klein, 2015; Lee et al., 2021). However, the strength of the relationship between risk perception and behavior varies situationally (Brewer et al., 2004; Bruine de Bruin & Bennett, 2020). Understanding links between shifts in risk perception and behavior engagement can help identify and recommend BIs that are aligned with population risk perceptions to or promote accurate risk messaging drive protective behavioral changes.

### 1.1. Study Setting

Centre County, PA is home to a large university that is surrounded by a non-student community in a suburban to rural setting (Fig. 1a). In 2020, the county had a population of 158,172 (U.S. Census Bureau, 2020) and with a full-time undergraduate student enrollment of approximately 40,600 (Office of planning, assessment, and institutional research, 2021) at the University Park campus in Centre County from the Fall of 2019. The non-student population is present throughout the year while the undergraduate student population fluctuates seasonally, corresponding to the academic calendar.

**Figure 1.**
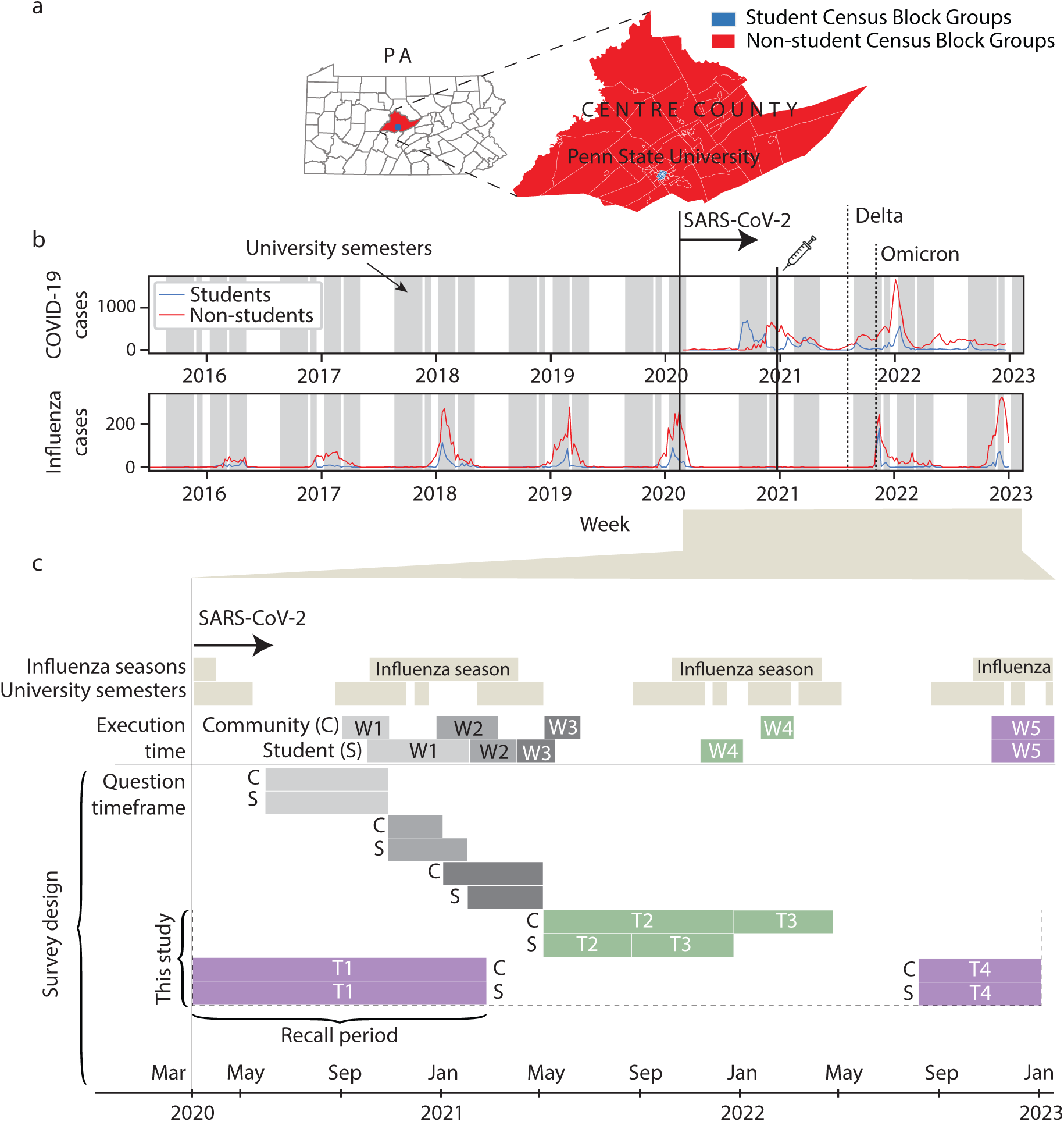
a) Census block groups in Centre County categorized by student (blue) and non-student (red) populations. b) Weekly COVID-19 and seasonal influenza incidence. The solid vertical line shows the local emergence of SARS-CoV-2 and dotted vertical lines show the regional emergences of Delta and Omicron variants. COVID-19 vaccinations in Centre County began in late December 2020. c) Timeframes for the Data4Action longitudinal survey among both cohorts. This study focused on behaviors and perceptions during timeframes T1-T4 that were measured during surveys W4 and W5; W4 targeted T2 and T3 while W5 targeted timeframes T1 and T4.

Based on the 2020 Census, census block groups (CBGs) for which >50% of ACS responses reported enrollment as a full time undergraduate student had a median age of 21. We defined these CBGs as student-dominated CBGs. The median age of all other CBGs in the county was in the 45-49 range and primarily represented the non-student population. Additionally, the median household income for student-dominated CBGs was between $25,000 and $29,999, while for all other CBGs, it was within the $60,000 to $74,999 range, based on the 5-year ACS adjusted for the inflation rate in 2020.

Early in the pandemic, both the student and non-student populations demonstrated high levels of adherence to the BIs that were suggested by the local government and university. Each cohort exhibited a distinct pattern of COVID-19 incidence (Bharti et al., 2022; Faust et al., 2021) (Fig. 1b). In the fall of 2020, the student population was only partially present and instruction was largely remote. Despite that, during that semester, student COVID-19 cases rose rapidly and infected a high proportion of the cohort, and non-student cases increased slowly and infected a smaller proportion of the cohort. Upon the return of in-person instruction and students to Centre County in the Spring of 2021, COVID-19 cases among students increased from near absent levels while cases among non-students continued in a smaller second wave. A smaller proportion of the population from each cohort was infected compared to the first wave of the COVID-19. In the Fall of 2021, both student and non-student cases increased, but student cases decreased promptly, while non-student cases grew to their highest numbers.

Globally, seasonal influenza significantly decreased during the first year of implemented COVID-19 interventions. Similarly, in Centre County, the incidence of seasonal influenza during the 2020-2021 season was completely disrupted. This reduction in influenza reporting occurred despite increased testing capacity and no significant changes in vaccination rates. Weather factors strongly correlated with influenza seasons, such as absolute humidity, were not significantly different from prior years (Fig. S1). This indicates that BIs implemented to mitigate COVID-19 reduced the transmission of seasonal influenza in Centre County.

We sought to understand student and non-student cohort responses to BIs during different phases of the COVID-19 pandemic in Centre County. We surveyed students and non-students in Centre County about their behaviors at various time points during the first three years of the COVID-19 pandemic (Fig. 1c). We conducted cluster analysis to identify cohort level behaviors that showed similar dynamics across these three years. To measure health behaviors and risk perceptions, we assessed responses to 22 behavioral items of a longitudinal survey that targeted four timeframes during the study period.

We found a consistent pattern of declining compliance with protective behaviors and increasing engagement with risky behaviors over time in both student and non-student cohorts. We observed minor increases in the levels of protective behaviors among non-students that were correlated with the emergence of the Omicron variant. Within each cohort, we identified behaviors that exhibited similar dynamics (uptake, persistence, waning). Behaviors were clustered based on engagement levels and variation. We also compared the dynamics of newly introduced COVID-19 interventions to traditionally promoted BIs and found that newly introduced COVID-19 BIs (e.g., facemask and controlling gatherings and events) likely impacted the dynamics of multiple respiratory pathogens. Finally, we explored potential links between changes in behaviors and changes in risk perceptions in both cohorts. Our results identified some explanatory power of risk perception on BIs. These findings can help 1) categorize BIs, which can provide some predictability about their dynamics, and 2) leverage risk perception for effective BI recommendations for outbreak management.

## 2. Methods

### 2.1. Recruitment and participation

We longitudinally surveyed student and non-student cohorts in Centre County Pennsylvania about their pandemic attitudes and behaviors, including their adoption of suggested BIs following the local emergence of SARS-CoV-2. We conducted five waves (W1-W5) of surveys from 2020 to 2023 for each cohort (Fig. 1c). To gauge interest in participation and collect demographic details, we initially distributed a pre-survey to residents of Centre County (Arnold et al., 2021). Students were recruited using a similar internal survey through cold-emails, word-of-mouth, online news stories, and campus postings. Non-student participants were eligible if they were ≥ 18 years old, not registered as full-time undergraduate students, currently residing in Centre County and planning to reside in Centre County until at least June 2021 (W3), fluent in English, and capable of providing their own consent. Eligible student participants were ≥ 18 years old, enrolled at the main university campus as full-time undergraduate students, and intending to reside in the County for the next year. The non-student community underwent a single recruitment stage from June through September 2020, while new students were recruited at various points throughout the survey period to compensate for attrition resulting from graduation.

We focused on survey responses to 22 items that concentrated on BIs. Among the 22 survey items analyzed here, only six were identical across all five waves of surveys. Therefore, we primarily used data that were collected in the final two waves (W4 and W5) of the survey for both student and non-student cohorts. These questions asked about current and retrospective behaviors. W1, W2, and W3 took place during the first year of the pandemic, while W4 and W5 occurred in the second and third years, respectively.

In W4 and W5 surveys, participants were asked about their behaviors, perceptions, and attitudes across different time periods, covering the entire three years that had elapsed since the local emergence of SARS-Cov-2. Specifically, questions covering periods T1 and T4 (Fig. 1c) were asked in W5 while T2 and T3 were asked in W4. To address potential recall bias for retrospective survey items, particularly for the T1 period (labeled as the recall period in figure 1c), we assessed the agreement between responses in W5 and the first three waves for the six survey items that were present in each of these waves.

### 2.2. Measuring behavior and risk perception

We surveyed participants on various BIs, including face masking, social distancing, personal hygiene behaviors (such as hand washing, not touching eyes, and covering nose and mouth when coughing or sneezing), attendance at gatherings of different sizes, attendance at concerts and sports events, attendance at indoor and outdoor gatherings, restaurant dining, and working or attending class remotely. For each of the 22 survey items, participants were asked, “How often did you follow the public health recommendations [about this behavior]?” during a specific time period. The survey offered participants five main response options: never, rarely, sometimes, most of the time, and always as well as “not applicable” and/or “prefer not to answer.”

We captured risk perception of participants associated with each behavior by asking, “To what extent do you associate any of the following with the risk of COVID-19 transmission or infection?” in two cross sections in time for each cohort. Participants were surveyed with five main response options: decreases safety a lot, decreases safety a little, does not impact safety, increases safety a little, and increases safety a lot, as well as “not applicable” and/or “prefer not to answer.”

### 2.3. Analyzing behavior and risk perception

We used the common ordered categories method, also known as Likert scale variables, to measure behaviors of participants and assess their perception of risk. In parametric methods, ordinal variables can be analyzed using equally spaced scores, Ridit scores (Bross, 1958), conditional median (Fernández et al., 2020), or conditional mean and scoring functions (Fielding, 1993). Our analysis of behaviors using Ridit scores revealed minimal differences in behavioral changes over time compared to the equal interval method (see section S3 and Fig. S2 in supplementary materials for a comprehensive comparison of results using equal interval and Ridit analysis scales). Given that Ridit analysis introduces uncertainties related to baseline population and considering the simplicity of the equal interval method, we have opted to present results using the latter approach.

**Figure 2.**
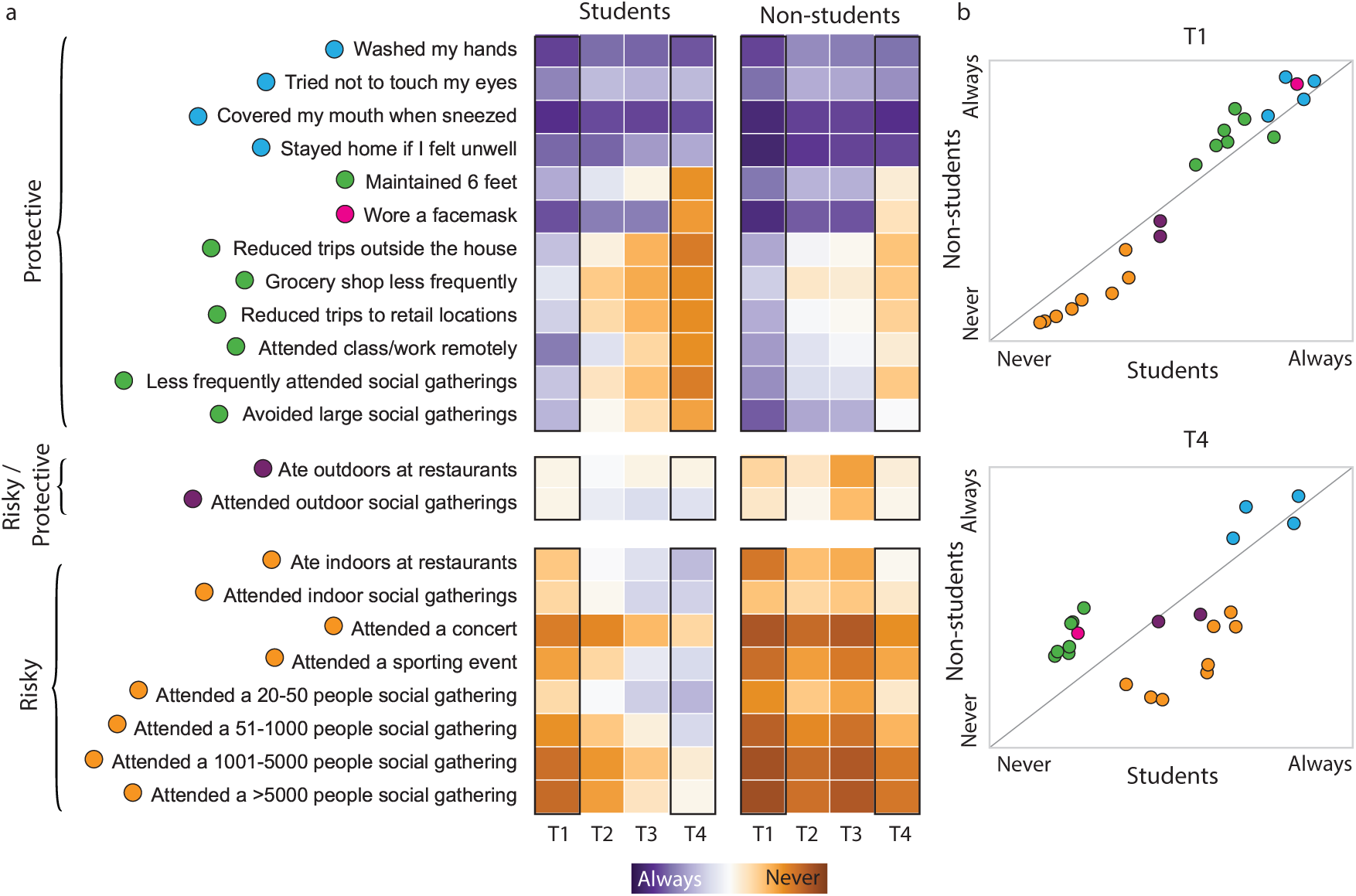
Pandemic related behaviors among student and non-student cohorts during four survey periods T1-T4 (responses collected during W4 and W5). A) Behavior levels during T1-T4. Both student and non-student populations showed trends of decreases in protective behaviors and increases in risky behaviors. B) Comparison of BIs between students and non-students at the onset (T1) and end (T4) of the study period color coded by clusters in Fig. 3.

We assigned values from 0 to 4 to both behavioral action and risk perception survey items, where 0 represented “never” or “decreases safety a lot,” and 4 represented “always” or “increases safety a lot.” Then, we calculated the average of individual responses for each survey item to represent the cohort level behavior or risk perception during each time period. Answers of “not applicable,” “prefer not to answer,” or no response were treated as if the respondent was not asked the survey item, resulting in different sample populations for each item.

We assessed the shifts in risk perceptions and behaviors for both cohorts between timeframes T3 and T4. To visualize the shifts in risk perception and potential links to shifts in behavior, we regressed behaviors against risk perception. We need to interpret the results cautiously due to inherent assumptions associated with parametrizing cohort level behaviors using the equal interval method.

### 2.4. Behavioral clusters

BIs that are interdependent in their use or effect may be promoted as a set and they may increase and decrease together (Smith et al., 2023). We used hierarchical clustering to identify clusters of behaviors for each cohort based on levels of engagement and variations in usage of BIs over time. We determined the number of clusters using silhouette scores and visual analysis.

### 2.5. Recall bias

Recall bias occurs when people inaccurately remember or misreport past events or experiences. In our study, responses of participants about the T1 period were subject to recall bias because they were collected approximately two years later, during W5. Because T1 spanned the first three waves of data collection, we assessed the agreement between estimated cohort level behaviors collected during the first three survey waves (W1-W3) and in W5. Cohort level behaviors were calculated as the average of reported behaviors from W1 to W3, weighted by the number of overlapped months with T1.

There are two approaches to comparing mean values of two cross-sectional sample populations. Paired samples are used when all participants are the same between the two samples and independent samples are employed when participants are not necessarily the same. Due to the static nature of the non-student cohort compared to the turnover of the student participants, we measured recall bias using both paired and independent sample methods with the non-parametric Wilcoxon signed-rank test and Mann-Whitney U test, respectively. These tests are distribution-independent and are used to determine the significance of recall bias in small sample populations.

## 3. Results

### 3.1. Data 4 Action survey

Out of 1530 eligible and willing to participate non-student cohort residents, 833 individuals participated in W4, and 684 individuals participated in W5. Among the 1297 eligible students who participated in at least one of the waves, 238 and 356 individuals participated in W4 and W5, respectively. In W1, the median student age was 21, the median non-student age was 52.5, the self-reported median household income for students was under $25k, and for non-students, it was $75k to $99,999. These statistics aligned with census extracted age and income from student- and non-student-dominated CBGs (CBGs do not precisely represent student and non-student populations). Additional detailed information on participation, demographics, and socioeconomic characteristics of the two cohorts in each wave is provided in Table S1 in the supplementary materials.

### 3.2. Uptake and waning of BIs

We categorized pandemic related behaviors as protective when they were intended to reduce transmission (e.g., wearing a facemask, working remotely) or risky when they could increase transmission (e.g., attending gatherings or events). Certain behaviors were not easily classified because their likelihood of reducing or increasing transmission was subject to an undefined comparison behavior. For example, outdoor restaurant dining could be compared to indoor restaurant dining, which is more risky, or not dining out at all, which is less risky. Similarly, outdoor social gatherings could be compared to riskier indoor social gatherings or not attending social gatherings.

BIs were introduced in 2020 and had peak levels of engagement in 2020 (T1). We found that both cohorts consistently decreased practicing protective behaviors and increased engagement with risky behaviors from T1 forward (Fig. 2a). In the non-student cohort, the regional emergence of the Omicron wave in November 2021 coincided with a slight decline in risky behaviors and outdoor activity (T3). This could represent a feedback loop between behavior and disease incidence.

Compared to students, non-students reported fewer changes in their practice of BIs over the study timeframes (Fig. 2b). When SARS-CoV-2 emerged locally, (T1) both cohorts acted similarly, with the non-student cohort adopting protective behaviors and avoiding risky behaviors at a slightly higher average value than the student cohort. As the pandemic continued, students reported earlier reductions in protective behaviors and increases in risky behaviors. These trajectories continued through T4.

Personal hygiene behaviors remained relatively stable throughout the study period in both cohorts. Outdoor activities, which we could not classify as risky or protective, showed consistency in T1 and T4 but displayed slight declines in T2 and T3 among non-students. T2 and T3 both included winter seasons, which can prohibit outdoor activities in central Pennsylvania. Other behaviors were monotonic in both student and non-student cohorts, such as masking, which was high at early timepoints and declined.

### 3.3. Behavior clusters

Using hierarchical clustering to explore the trajectories of BIs from T1 to T4, we found the optimal number of clusters for both student (triangles) and non-student (circles) cohorts was four, as determined by the silhouette score (Fig. 3). Two differences between student and non-student populations were 1) among students, wearing a facemask formed one distinct class, and 2) non-students exhibited a separate class associated with outdoor activities, namely eating outdoors at restaurants, and attending outdoor social gatherings.

**Figure 3.**
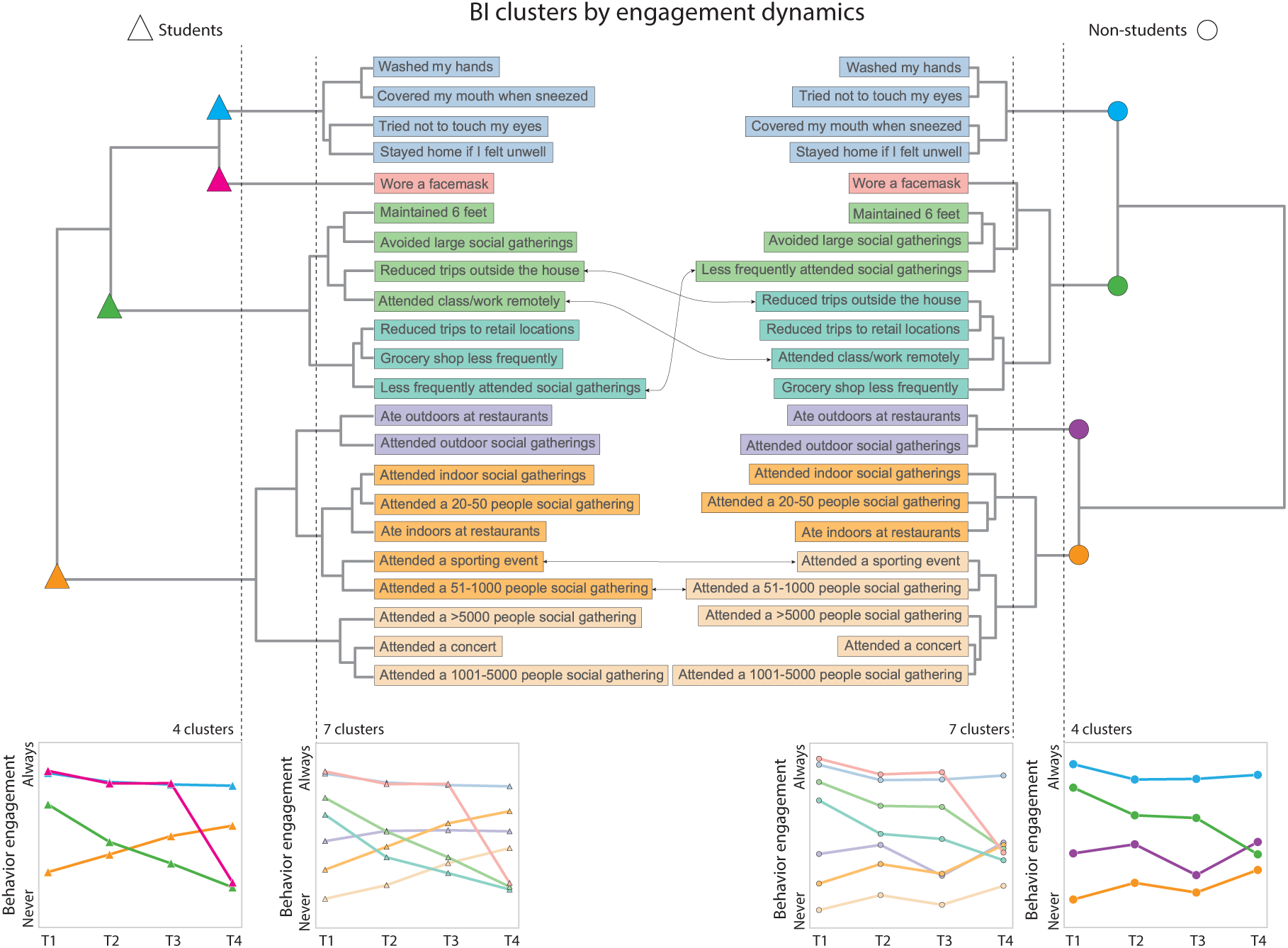
BI clusters by engagement levels and variation over time display similarities between the two cohorts under four-cluster (based on silhouette score) and seven-cluster cut-offs.

To delve deeper into behavior clusters, we chose a lower cut-off value on the dendrogram, resulting in seven clusters for each cohort. We observed almost consistent clusters among both student and non-student cohorts. With seven clusters, wearing a facemask formed a distinct cluster for both cohorts. Gatherings and events, including small and large ones as well as indoor/outdoor group activities, social distancing, and trips also formed distinct clusters. However, we observed some BIs interchange when comparing the clusters from the student and non-student cohorts (arrows in Fig 3.).

Overall, students consistently decreased their reduction of trips outside the house and remote class attendance while increasing attendance at gatherings and events (Fig. 3, line plots below). Non-student behavior clusters exhibited trends that may indicate more caution, with increases in protective BIs coinciding with the local emergence of the Omicron variant during the T3 survey for this cohort. Notably, personal hygiene behaviors and staying home when feeling ill maintained consistent, high levels for both cohorts. Similarly, outdoor dining and attending outdoor gatherings and events remained stable with a moderate level of engagement. The use of face masks declined significantly after T3 in both cohorts.

### 3.4. Risk perception and behavior

We analyzed links between the perception of risk and behavioral interventions. We measured both at the time of data collection and the self-reported practice of that behavior at the same time. We captured risk perception in W4 and W5.

For most behaviors surveyed, the risk perception and behavior engagement changed in the same direction over time (increase or decrease) (Fig. 4). Within the student cohort (Fig. 4, left panel), the greatest change in risk perception was associated with the frequency of attending gatherings and events and the size of gatherings and events, while the least change was associated with eating outdoors at restaurants, followed by personal hygiene behaviors. In the non-student population (Fig. 4, center panel), we observed similarly large changes in the perception of risk associated with the frequency of attending and the size of gatherings.

**Figure 4.**
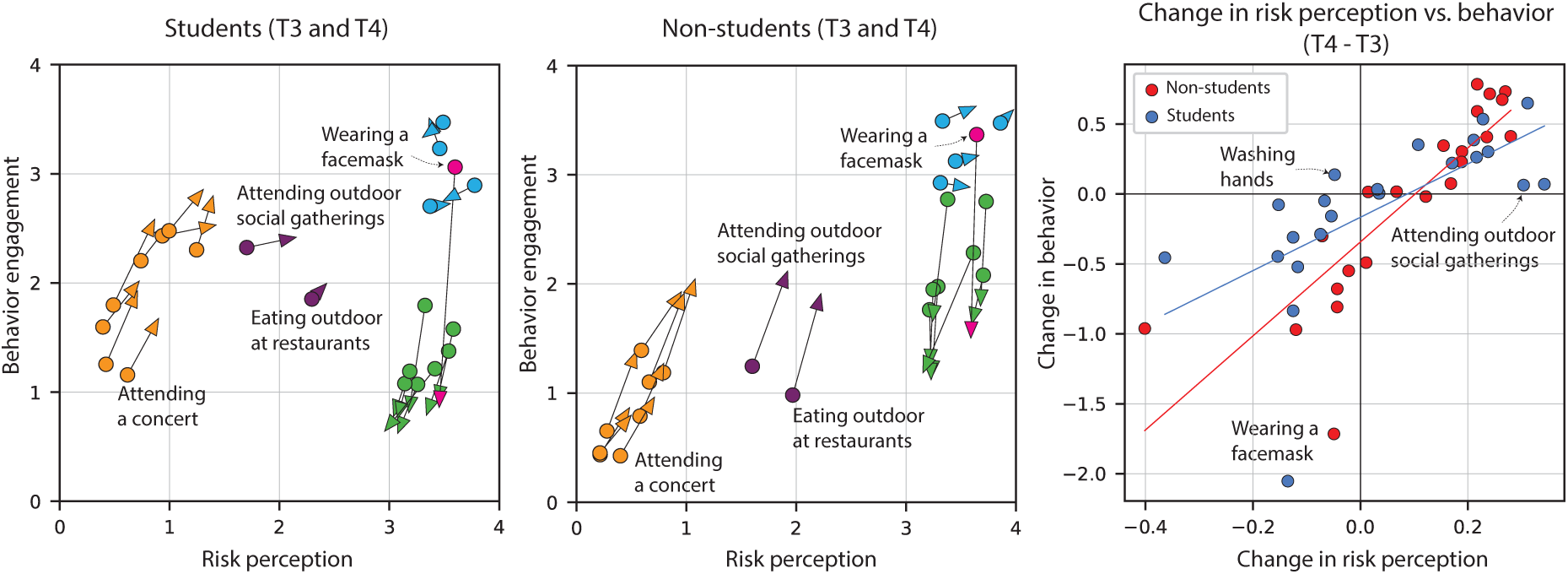
Risk perception and behavior during survey periods T3 and T4. Left, center: for each cohort, each arrow base represents the risk perception and level of engagement with a behavior during T3, and each arrowhead indicates the value during the T4 period. On each axis, a high value (4) indicates safety or action while a low value (0) indicates no action or safety. Right: the change in behavior against the change in risk perception for both cohorts (T4 minus T3).

We assessed the link between the change in risk perception and the change in BIs among both cohorts (Fig. 4, right panel). The ordinary least squares regression analysis for the student cohort revealed an R-squared of 0.416, implying that approximately 41.6% of the change in BIs was explained by change in risk perceptions. The R-squared for the non-student community regression was 0.70, indicating that changes in risk perceptions may explain even more about changes in BIs for this cohort. We remain cautious about these statistics since ordinal variables were transformed to variables with equal interval digits.

BIs with large changes in risk perception and action (trips, gatherings and attending work/class remotely) demonstrated that perceptions could change quickly and give rise to rapid changes in behaviors. Personal hygiene and staying home if feeling unwell remained stable in both risk perception and engagement level. However, it was notable that both cohorts perceived masking as a behavior that greatly increases safety with minimal change in perception between the two time points while reporting a significant reduction in masking behavior. This indicates masking behavior was heavily impacted by factors other than risk perception.

### 3.5. Recall bias

Across W1-W5, we asked individuals about their actions with respect to a subset of six protective BIs (washing hands, wearing a facemask, trying not to touch eyes, covering mouth when sneezed, staying home if feeling unwell, maintaining 6 feet distance, and avoiding crowded places). Based on the methodology explained in section 2.7, we used the responses to these six questions collected from the first three waves of the survey (W1-W3), which took place during the first year of the pandemic, to calculate the recall bias of these six behaviors from retrospective questions asked in W5.

We find recall bias is statistically significant for almost all these behaviors. The mean differences indicated that non-students tended to overstate their prior compliance to most of these six BIs whereas students tended to understate their prior compliance with these BIs. Among these six behaviors, we anticipate no significant impact on our results for wearing facemasks, washing hands, avoiding touching eyes, covering the mouth when sneezing, and maintaining six feet (among non-students) since the average recall error was small. For maintaining six feet among students, the average recall error was large, indicating that students understated their compliance in T1. However, we confirmed that this discrepancy did not alter clusters of BIs.

Recall bias for staying home when feeling unwell could have significant implications for our results. This behavior was understated by both cohorts in T1, with large average recall errors for students and non-students. That means both cohorts stayed home when sick during T1 more than in subsequent timeframes, resulting in a less stable behavior dynamic. For the student cohort, we are cautious when interpreting the average differences, as they are based on a small population size (n=37). A detailed analysis of recall bias is in section S3 of the supplementary materials.

## 4. Discussion

Across the first three years of the COVID-19 pandemic, both student and non-student populations in Centre County showed a trend of declining engagement with protective behaviors and increasing engagement with risky behaviors. However, non-students displayed smaller change in BIs than students, and the emergence of the Omicron variant coincided with a slight decline in risky behaviors among the non-students.

Previous research showed that the novel behaviors introduced in Centre County in response to COVID-19 (e.g., masking, physical distancing, avoiding crowds) and traditional symptom-management behaviors that are reinforced annually during flu season (covering coughs and sneezes, staying home if ill) appear as packages of adopted behaviors (Smith, Small, et al., 2023). Our findings add to that by demonstrating tendencies of behaviors to cluster not only based on adoption but their level of engagement and change over time. Behaviors that were first introduced after the emergence of the COVID-19 pandemic were found in multiple separate clusters and distinct from the BIs that are traditionally promoted. Among students and non-students, clusters were largely composed of similar behaviors.

Cluster dynamics showed differences, which can help identify behaviors that most likely contributed to the massive disruptions in influenza incidence during the 2020-2021 season, seen as an absence of cases. The following two influenza seasons experienced dynamics that were similar to pre-pandemic seasonal influenza (Fig. 1b). BIs were the primary factors that significantly changed during the 2020-2021 season only. Among implemented BIs, behaviors including personal hygiene behavior and outdoor activities (dining and gatherings) remained stable during the study period for both cohorts (Fig. 3). This indicates that the stable behaviors mentioned, despite high levels of engagement, were not the primary contributors to the observed disruptions in seasonal influenza during 2020-2021. Instead, it is more likely that BI clusters with high levels of engagement early in the pandemic followed by waning levels, such as masking, trips outside the house, remote work and class attendance, and indoor gatherings, were important factors that influenced disruptions in seasonal influenza transmission.

A national survey completed before the emergence of SARS-CoV-2 showed that 55% of adult workers in America went to work when they were feeling sick ‘most of the time’ or ‘always’ (NPR, 2016). In Centre County, following the emergence of COVID-19, 89.79% of non-students and 63.72% of students in our survey responded that they stayed home when feeling sick ‘most of the time’ or ‘always.’ Although out data collected during W4 and W5 showed staying home when feeling ill displayed stable dynamics from T1 to T4, our recall bias analysis showed that participants reported practicing this behavior during T1 significantly more than during subsequent time frames. Together, this suggests that ‘staying home when feeling ill’ in Centre County was likely highest during T1 and higher during T2-T4 than before the pandemic. It is likely that more Centre County residents stayed home when feeling sick after the emergence of SARS-CoV-2 than before and that, after the first year, they maintained a slightly reduced form of that behavior. Staying home when feeling ill could potentially have played a role in disrupting seasonal influenza during the first year of COVID-19.

Importantly, this does not imply that traditionally promoted hygiene practices have no impact on respiratory disease transmission. Our methods cannot measure the impact of BIs that are consistently in place until there is a significant change in their level of compliance.

Our analysis showed a strong correlation between changes in risk perception and engagement levels for behavioral interventions. Changes in risk perception explained changes in BI more strongly among non-students than among students. When examining specific behaviors, we categorize the links between perception and action into three groups. In the first category, both behavior and risk perception change, either in the same or opposite directions. For instance, attending gatherings and events in both cohorts, as well as outdoor events among non-students, showed changes in behavior and risk perception in the same direction. We found no instances of both behavior and risk perception changing significantly and in opposite directions within this category.

In the second category, either the behavior or risk perception changes, while the other factor remains relatively stable or experiences minimal change. For example, wearing facemasks and, to a lesser extent, maintaining a 6-feet distance, taking trips outside the house, and attending classes or working remotely all changed while the associated risk perception did not change. This suggests that factors beyond risk perception, such as fatigue from practicing BIs, vaccination, or institutional policies, may have exerted significant influence on behavior. T4 occurred in the third year of the pandemic when a large proportion of Centre County’s population was vaccinated and the university relaxed facemask requirements and held in person classes. The third category encompasses instances where both behavior and risk perception do not change. This category included traditional personal hygiene BIs and dining outdoors at restaurants.

Understanding the relationship between risk perception and behavior can help tailor recommendations. For example, basic hygiene BIs are generally taught to children and heavily reinforced every influenza season while novel BIs, such as wearing facemasks, were only recently promoted and can change quickly. A well-timed informational campaign could focus on the risks associated with respiratory viruses and the benefits of wearing a mask to reduce seasonal influenza transmission early in flu season.

Our study faced several limitations. First, responses regarding the T1 period that were collected during W5 were susceptible to recall bias. Additionally, the turnover in the student cohort throughout the study prevented us from studying a true longitudinal cohort. Although cohort surveys were rolled out in parallel, student and non-student cohorts were not always surveyed across identical time frames (T2 and T3). Even small offsets meant that one cohort might be more aware of an emerging novel variant or a new BI recommendation during the rapidly changing global pandemic. The statistical analysis of Likert score variables, especially with a high number of variables and categories, presented challenges. We employed the equal interval parametric method to interpret population-level results, necessitating caution in interpreting the statistical findings. Our survey began after the emergence of COVID-19, limiting our ability to accurately measure behaviors that preceded COVID-19, which would provide insights into the full range of variations of traditionally promoted behaviors.

Future studies could follow the behavior of these cohorts over an extended timeframe. Specifically, they could explore whether behaviors that remained stable during this study might undergo changes in the future (e.g., change in staying home when feeling unwell or personal hygiene behavior). Monitoring the levels of these behaviors and tracking them through perturbations, such as future emerging respiratory pathogens, could provide valuable insights.

## 5. Conclusions

This study explored BIs, their dynamics, patterns of clustering by changes in engagement levels, and the association between changes in risk perception and behavior among students and non-students within Centre County, Pennsylvania. We focused on behaviors that were targeted by pandemic BIs, highlighting a wide range of interventions such as longstanding personal hygiene practices and behaviors that were introduced in response to COVID-19. Our findings indicate BIs waned over time with some cohort specific fluctuations, potentially in response to COVID-19 dynamics. Additionally, we observed a significant tendency for behaviors to cluster based on engagement levels and variations in those levels. Our results underscored the explanatory power of risk perception for pandemic-related behaviors, which was different between the two cohorts studied. Categorizing behaviors into clusters based on their temporal variations and examining their relationship with risk perception offers a systematic approach to understanding behavioral dynamics and facilitates the design of effective interventions.

## Data Availability

All data produced in the present study are either publicly available with links provided or available upon reasonable request to the authors.

https://github.com/bhartilab/behavioral_interventions/tree/main

## Acknowledgements

We thank Brian Lambert for contributing to data retrieval and technical problem solving. We also thank the Pennsylvania Department of Health for providing access to Influenza incidence data.

## Author contributions

NB designed the study. NB and CE developed the survey instrument and collected the data for W4 and W5. NB and BN conceptualized the methodology. BN conducted the analyses. BN wrote the first draft of the manuscript with input from NB. All authors contributed to writing and editing the final draft.

## Ethics and Data availability statement

The Data4Action study received approval from Penn State University’s Institutional Review Board (STUDY00015547).

Academic researchers must register to receive access to SafeGraph data at no charge for non-commercial purposes. SafeGraph data are anonymized and aggregated. Our work with the data provided by SafeGraph did not qualify as human subjects research as defined by The Pennsylvania State University’s IRB. SafeGraph approved the use of their data for this study.

No administrative permissions were required to obtain COVID-19 incidence data; datasets used were freely available in the public domain here: https://data.pa.gov/api/views/j72v-r42c/rows.csv?accessType=DOWNLOAD, and https://virusinfo.psu.edu/covid-19-dashboard/. Influenza incidence data for non-students were provided by PA DOH and can be requested directly from them.

Census block group data are available here: https://www.census.gov/data/developers/data-sets/acs-5year.html.

Aggregate summaries of the Data4Action survey data and code to reproduce figures are available on GitHub (https://github.com/bhartilab/behavioral_interventions/tree/main). Additional data can be made available upon reasonable request to the corresponding author.

## Supplementary materials

### S1. Absolute humidity and foot traffic

Figure S1 illustrates the weekly absolute humidity (*g*/*m*^3^), and the number of visitors to places of interest from SafeGraph data, which includes specific physical locations like businesses, parks, stores, or other venues. The relative number of visitors is commonly utilized as a metric for measuring ‘staying at home’ (Kang et al., 2020). The number of visitors in both cohorts gradually increased after a significant initial drop at the onset of the pandemic, returning to pre-pandemic levels by late 2021. Weather data are from the Automated Surface Observing Systems (ASOS) (https://www.weather.gov/asos/) and we used the riem package in R (https://cran.r-project.org/web/packages/riem/index.html) to calculate absolute humidity.

**Figure S1.**
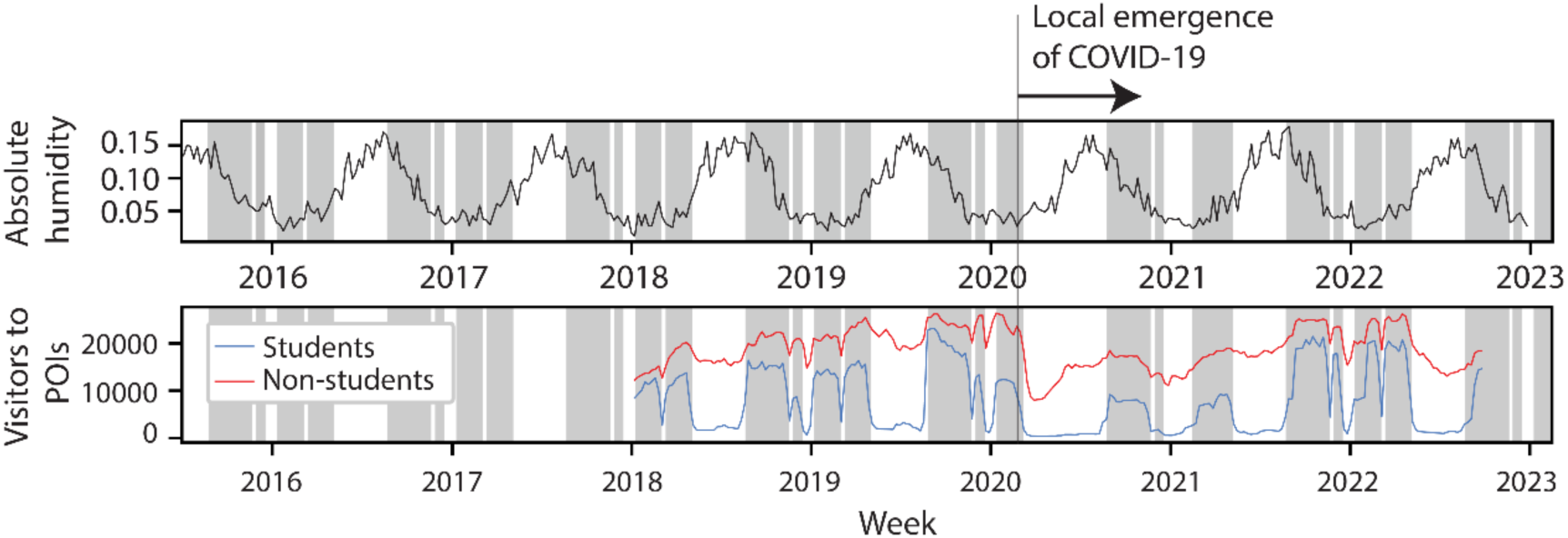
For Centre County, absolute humidity (top) and the number of student and non-student visitors to points of interest (POIs), derived from mobile device data (bottom).

### S2. Descriptive statistics

**Table S1.** Demographic and socioeconomic characteristics of student and non-student participants in all waves.

|  | W1 |  | W2 |  | W3 |  | W4 |  | W5 |  |
| --- | --- | --- | --- | --- | --- | --- | --- | --- | --- | --- |
|  | Students<br>(N=884) | Non-students<br>(N=974) | Students<br>(N=559) | Non-students<br>(N=905) | Students<br>(N=481) | Non-students<br>(N=867) | Students<br>(N=238) | Non-students<br>(N=833) | Students<br>(N=356) | Non-students<br>(N=684) |
| **Execution** |  |  |  |  |  |  |  |  |  |  |
| Start date | 10/24/2020 | 8/31/2020 | 1/27/2021 | 12/8/2020 | 4/2/2021 | 5/6/2021 | 11/3/2021 | 1/26/2022 | 10/26/2022 | 10/26/2022 |
| End date | 02/28/2021 | 12/3/2020 | 3/4/2021 | 3/5/2021 | 5/5/2021 | 6/18/2021 | 12/14/2021 | 4/29/2022 | 12/31/2022 | 12/31/2022 |
| **Age** |  |  |  |  |  |  |  |  |  |  |
| Median | 21 | 52.5 | 21 | 53 | 21 | 54 | 19 | 51 | 18 | 52 |
| Min | 18 | 18 | 18 | 19 | 18 | 19 | 17 | 18 | 18 | 18 |
| Max | 67 | 99 | 47 | 99 | 67 | 99 | 35 | 89 | 32 | 99 |
| Missing | 3 (0.3%) | 0 | 3 (0.5%) | 0 | 2 (0.4%) | 0 | 6 (2.5%) | 1 (0.1%) | 43 (12.1%) | 0 |
| **Gender** |  |  |  |  |  |  |  |  |  |  |
| Male | 295<br>(33.4%) | 297<br>(30.5%) | 188<br>(33.6%) | 277<br>(30.6%) | 161<br>(33.5%) | 268<br>(31.0%) | 72 (30.3%) | 268<br>(32.2%) | 105<br>(29.5%) | 217<br>(31.7%) |
| Intersex | 1 (0.1%) | 0 | 0 | 0 | 1 (0.2%) | 0 | 1 (0.4%) | 0 | 1 (0.3%) | 0 |
| Female | 583<br>(66.0%) | 671<br>(68.9%) | 359<br>(64.2%) | 623<br>(68.8%) | 311<br>(64.7%) | 596<br>(68.7%) | 153<br>(64.3%) | 564<br>(67.7%) | 223<br>(62.6%) | 466<br>(68.1%) |
| Prefer not to<br>answer | 0 | 0 | 0 | 0 | 0 | 0 | 0 | 0 | 0 | 0 |
| Missing | 5 (0.6%) | 2 (0.2%) | 11 (2.0%) | 2 (0.2%) | 8 (1.7%) | 2 (0.2%) | 12 (5%) | 1 (0.1%) | 27 (7.6%) | 1 (0.1%) |

Table S1. Demographic and socioeconomic characteristics of student and non-student participants in all waves (cont.).
|  | W1 |  | W2 |  | W3 |  | W4 |  | W5 |  |
| --- | --- | --- | --- | --- | --- | --- | --- | --- | --- | --- |
|  | Students<br>(N=884) | Non-students<br>(N=974) | Students<br>(N=559) | Non-students<br>(N=905) | Students<br>(N=481) | Non-students<br>(N=867) | Students<br>(N=238) | Non-students<br>(N=833) | Students<br>(N=356) | Non-students<br>(N=684) |
| **Race** |  |  |  |  |  |  |  |  |  |  |
| White | 717<br>(81.1%) | 863<br>(88.6%) | 458<br>(81.9%) | 806<br>(89.1%) | 398<br>(82.7%) | 780<br>(90.0%) | 180<br>(75.6%) | 768<br>(92.2%) | 241<br>(67.7%) | 630<br>(92.1%) |
| Other races | 114<br>(12.9%) | 60<br>(6.2%) | 61<br>(10.9%) | 51 (5.6%) | 52<br>(10.8%) | 40<br>(4.6%) | 32<br>(13.4%) | 35<br>(4.2%) | 55<br>(15.4%) | 25 (3.7%) |
| More than<br>one race | 36 (4.1%) | 39<br>(4.0%) | 25 (4.5%) | 38 (4.2%) | 19 (4.0%) | 35<br>(4.0%) | 12 (5.0%) | 24<br>(2.9%) | 19 (5.3%) | 20 (2.9%) |
| Prefer not to<br>answer | 5 (0.6%) | 11<br>(1.1%) | 2 (0.4%) | 9 (1.0%) | 2 (0.4%) | 9 (1.0%) | 2 (0.8%) | 4 (0.5%) | 10 (2.8%) | 7 (1%) |
| Missing | 12 (1.4%) | 1 (0.1%) | 13 (2.3%) | 1 (0.1%) | 10 (2.1%) | 2 (0.2%) | 12 (5%) | 2 (0.2%) | 10 (2.1%) | 2 (0.3%) |
| **Education** |  |  |  |  |  |  |  |  |  |  |
| High School<br>Diploma/<br>GED or less | NA | 20<br>(2.1%) | NA | 17 (1.9%) | NA | 14<br>(1.6%) | NA | 15<br>(1.8%) | NA | 12 (1.8%) |
| Associate<br>degree/vocat<br>ional<br>Training/So<br>me college | NA | 103<br>(10.6%) | NA | 93<br>(10.2%) | NA | 83<br>(9.6%) | NA | 67 (8%) | NA | 56 (8.2%) |
| Bachelor's<br>degree | NA | 289<br>(29.7%) | NA | 263<br>(29.1%) | NA | 244<br>(28.1%) | NA | 231<br>(27.7%) | NA | 194<br>(28.4%) |

Table S1. Demographic and socioeconomic characteristics of student and non-student participants in all waves (cont.).
|  | W1 |  | W2 |  | W3 |  | W4 |  | W5 |  |
| --- | --- | --- | --- | --- | --- | --- | --- | --- | --- | --- |
|  | Students<br>(N=884) | Non-students<br>(N=974) | Students<br>(N=559) | Non-students<br>(N=905) | Students<br>(N=481) | Non-students<br>(N=867) | Students<br>(N=238) | Non-students<br>(N=833) | Students<br>(N=356) | Non-students<br>(N=684) |
| Graduate/Professional Degree | NA | 559<br>(57.4%) | NA | 516<br>(57.0%) | NA | 490<br>(56.5%) | NA | 494<br>(59.3%) | NA | 402<br>(58.8%) |
| Prefer not to answer | NA | 0 | NA | 0 | NA | 0 | NA | 0 | NA | 0 |
| Missing | NA | 3 (0.3%) | NA | 16 (1.8%) | NA | 36 (4.2%) | NA | 26 (3.1%) | NA | 20 (2.9%) |
| **Income** |  |  |  |  |  |  |  |  |  |  |
| Under \$25k | NA | 87 (8.9%) | NA | 77 (8.5%) | NA | 71 (8.2%) | 193<br>(81.1%) | 70 (8.4%) | 269 (75.6%) | 54 (7.9%) |
| \$25k to \$49,999 | NA | 186<br>(19.1%) | NA | 181 (20%) | NA | 171<br>(19.7%) | 3 (1.3%) | 147<br>(17.6%) | 5 (1.4%) | 121<br>(17.7%) |
| \$50k to \$74,999 | NA | 187<br>(19.2%) | NA | 175<br>(19.3%) | NA | 164<br>(18.9%) | 2 (0.8%) | 186<br>(22.3%) | 3 (0.8%) | 150<br>(21.9%) |
| \$75k to \$99,999 | NA | 143<br>(14.7%) | NA | 134<br>(14.8%) | NA | 131<br>(15.1%) | 0 | 130<br>(15.6%) | 7 (2%) | 101<br>(14.8%) |
| \$100k and over | NA | 196<br>(20.1%) | NA | 191<br>(21.1%) | NA | 191<br>(22.0%) | 1 (0.4%) | 239<br>(28.7%) | 3 (0.8%) | 206<br>(30.1%) |
| Prefer not to answer | NA | 85 (8.7%) | NA | 69 (7.6%) | NA | 67 (7.7%) | 35 (14.7%) | 59 (7.1%) | 65 (18.2%) | 51 (7.5%) |
| Missing | NA | 0 | NA | 0 | NA | 0 | 4 (1.7%) | 1 (0.1%) | 4 (1.1%) | 0 |

### S3. Equal interval and Ridit score analysis

Survey items we analyzed in the study have response options with ordered categories (always, most of the times, sometimes, rarely, never, and not applicable), also known as Likert scale variables. Ordinal variables can be treated like interval variables by assigning equally spaced scores, allowing the use of parametric statistics (e.g., population mean). In practice, the dissimilarity between adjacent levels of ordinal variables may vary. For example, the difference in compliance with wearing a facemask from ‘never’ to ‘rarely’ may be different than the difference between ‘most of the time’ and ‘always.’ Several methods address this issue, including Ridit scores (Bross, 1958), conditional median (Fernández et al., 2020), conditional mean and scoring functions (Fielding, 1993). However, these methods have their own shortcomings.

The Ridit scores method replaces category levels with scores calculated as the proportion of the sample population in lower categories plus one-half the proportion of the sample population in the category itself (Bross, 1958). These scores are then used to calculate statistical metrics. However, Ridit scores are limited by the need for an appropriate baseline population and can only offer a relative assessment of the measured variable. Due to changes in population size and participants in our study, Ridit scores are not considered the best choice.

Here we compare the Ridit scores and a statistic called mean Ridit with the average behavior from equal interval scaling for six behaviors. Ridit scores are relative to an empirical sample distribution. In our study, each wave constituted a sample population, and the surveyed individuals varied from wave to wave, with attrition among non-students and attrition and replacement among students. One wave is selected as the baseline population. Ridit scores are computed for this baseline population in both cohorts. These Ridit scores serve as a basis for calculating the weighted mean Ridit scores for behaviors observed in subsequent waves. The choice of an appropriate baseline population is crucial as it forms the reference against which other sets of individuals are compared. The baseline population should be a representative sample, capturing existing heterogeneities in the population. Given our focus on data collected during W4 and W5, we opt for a baseline timeframe within T1-T4. The most suitable baseline in our study is either T3 or T4 due to the lower likelihood of recall errors. We selected T3 and six variables common across all waves for comparison of population level statistics derived from equal interval and average Ridit scales.

Figure S2 compares the 6 behaviors dynamics from Ridit scores and equal interval for both cohorts. Figures S2a and S2c depict average population behavior obtained through equal interval scaling. The first three timeframes represent data collected from W1-W3, while the last three (T2-T4) represent data collected during W4 and W5. Figures S2b and S2d illustrate the average behavior derived from Ridit analysis. The baseline (T3) is indicated by a vertical dashed line. The mean Ridit score for the baseline population is defined as 0.5 (represented by horizontal dashed lines). This means that two randomly chosen students from time frame T3 are equally likely to wear masks.

**Figure S2.**
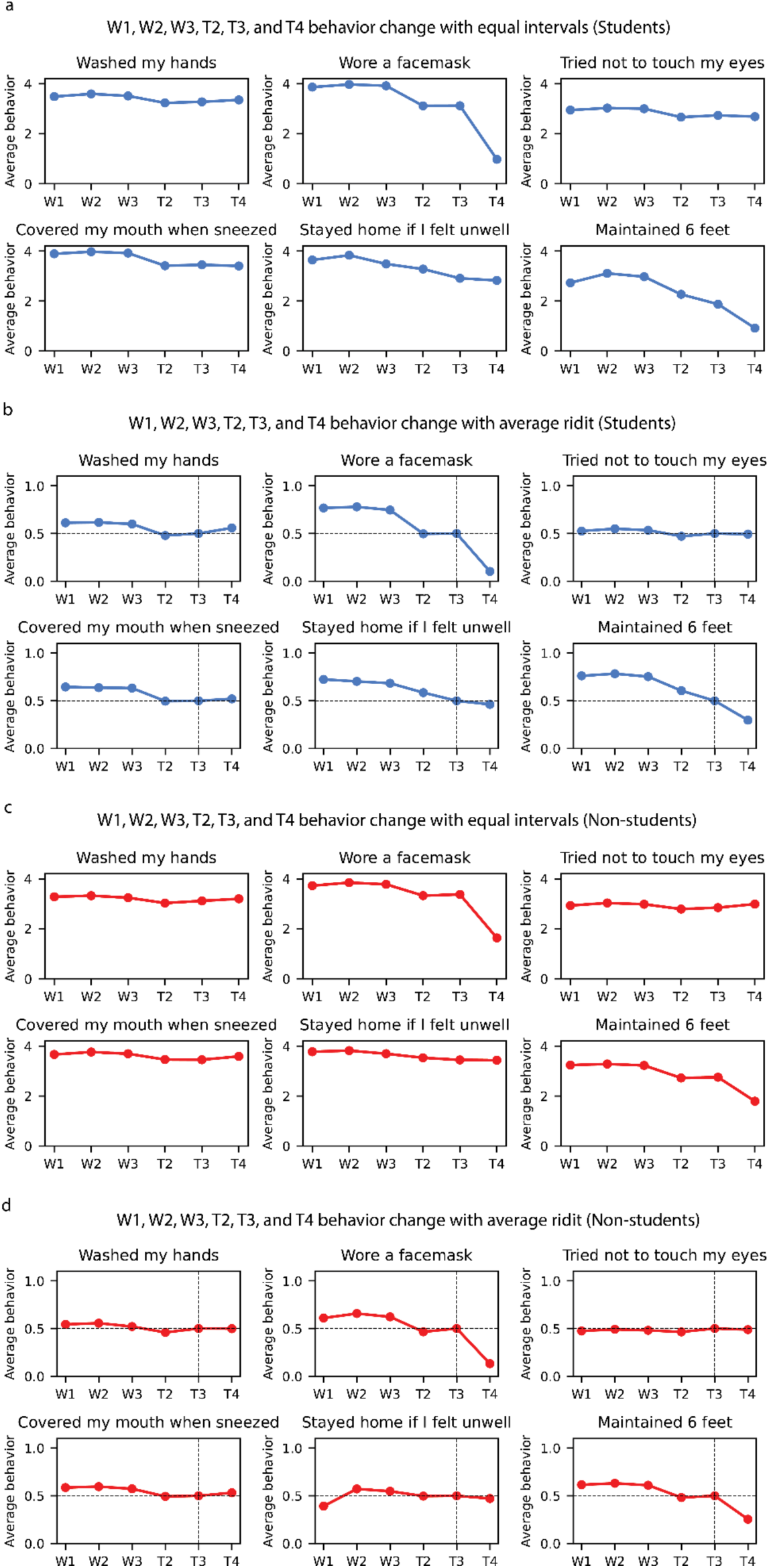
Comparison of average population behavior using equal interval versus Ridit analysis scaling; students in blue, non-students in red. a) Average student behavior derived from equal interval scaling. b) Average student behavior derived from Ridit score scaling. c) Average non-student behavior derived from equal interval scaling. d) Average non-student behavior derived from Ridit score scaling. Vertical dashed lines represent the baseline time frame (T3) for Ridit analyses. Both methods show similar dynamics in behaviors.

For example, a mean Ridit of 0.78 for students wearing a facemask in W2 indicates that the probability of an individual wearing a mask in W2 (early in the pandemic) is estimated to be 0.78 compared to an individual in time frame T3 (later in the pandemic) with probability of 0.5.

Visual comparison of the two methods reveals minimal differences in their dynamics. Consequently, we have chosen to present results using equal interval scaling, which is easily communicable and avoids the limitations associated with selecting a baseline population.

### S4. Recall bias

Across W1-W5, we asked individuals about their behaviors with respect to a subset of six protective BIs (washing hands, wearing a facemask, trying not to touch eyes, covering mouth when sneezing, staying home when unwell, maintaining 6 feet distance, and avoiding crowded places). We used responses to these six questions in the first three waves (first year of the pandemic) to calculate the recall bias of these six behaviors in W5.

Responses from participants were subject to potential recall biases regarding the T1 period, which were collected approximately two years later, during W5. We assessed the agreement between estimated cohort level behaviors collected during the first three waves and the retrospective responses collected in W5. Because the timeframe included in T1 spanned the first three waves of survey data collection, cohort level behaviors were calculated as the average of reported behaviors from W1 to W3, weighted by the number of months overlapping with T1.

From the non-student cohort surveys, we analyzed a subset of individuals who participated in W1, W2, W3, and W5 (N=444) and evaluated the agreement between sample average behavior about T1 that was reported in W5 and the responses collected in real time in W1-W3 using paired sample t-test statistics. We used paired sample t-test statistics because the individual subset demonstrated no significant difference in average behavior compared to the entire participant sample in each wave.

To calculate recall bias, retrospective responses about each of these six behaviors in W5 was statistically compared to the weighted average of behaviors in W1, W2, and W3, weighted by the number of overlapping months between surveys W1, W2, and W3 and the retrospective time period asked about in W5. Results showed that the recall bias was statistically significant for four behaviors (washing hands, trying not to touch eyes, staying home when unwell, and maintaining 6 feet distance). Among these four behaviors, the mean differences indicated that non-student members tended to understate their compliance with staying home if they felt unwell and overstate their compliance with the other three behaviors (Table S2 and Fig. S3).

For the student surveys, only a small number of individuals participated in W1, W2, W3, and W5 (n=37). For this cohort, we used independent t-test statistics to measure the population-wide recall bias among students by considering the entire sample of participants in each wave to evaluate agreement in average population-level behaviors surveyed during and after the first year of the pandemic.

**Table S2.** Recall bias regarding behavior during T1. No bias means that responses about T1 from W5 were in agreement with weighted average behavior responses from W1, W2, and W3. Negative (-) bias means participants understated their prior compliance, and positive (+) bias means they overstated their prior compliance when asked about retrospective behaviors during W5.

| Behavior | Paired samples (Wilcoxon signed-rank test) |  |  |  | Independent samples (Mann-Whitney U test) |  |  |  |
| --- | --- | --- | --- | --- | --- | --- | --- | --- |
|  | Students (N=37) |  | Non-students (N=444) |  | Students (N1=451, N2=345) |  | Non-students (N1=815, N2=650) |  |
|  | Bias direction | p-value | Bias direction | p-value | Bias direction | p-value | Bias direction | p-value |
| Washed hands | — | ns | + | *** | — | ns | + | *** |
| Wore a facemask | — | *** | — | . | — | *** | — | ns |
| Tried not to touch eyes | + | ns | + | *** | + | *** | + | *** |
| Covered mouth when sneezed | — | ns | + | ns | — | * | — | *** |
| Stayed home when feeling unwell | — | *** | — | *** | — | *** | — | *** |
| Maintained 6 feet distance | — | * | + | * | — | *** | — | *** |
| Significance codes: 0.001 (***), 0.01 (**), 0.05 (*), 0.1 (.), non-significant (ns). |  |  |  |  |  |  |  |  |

**Figure S3.**
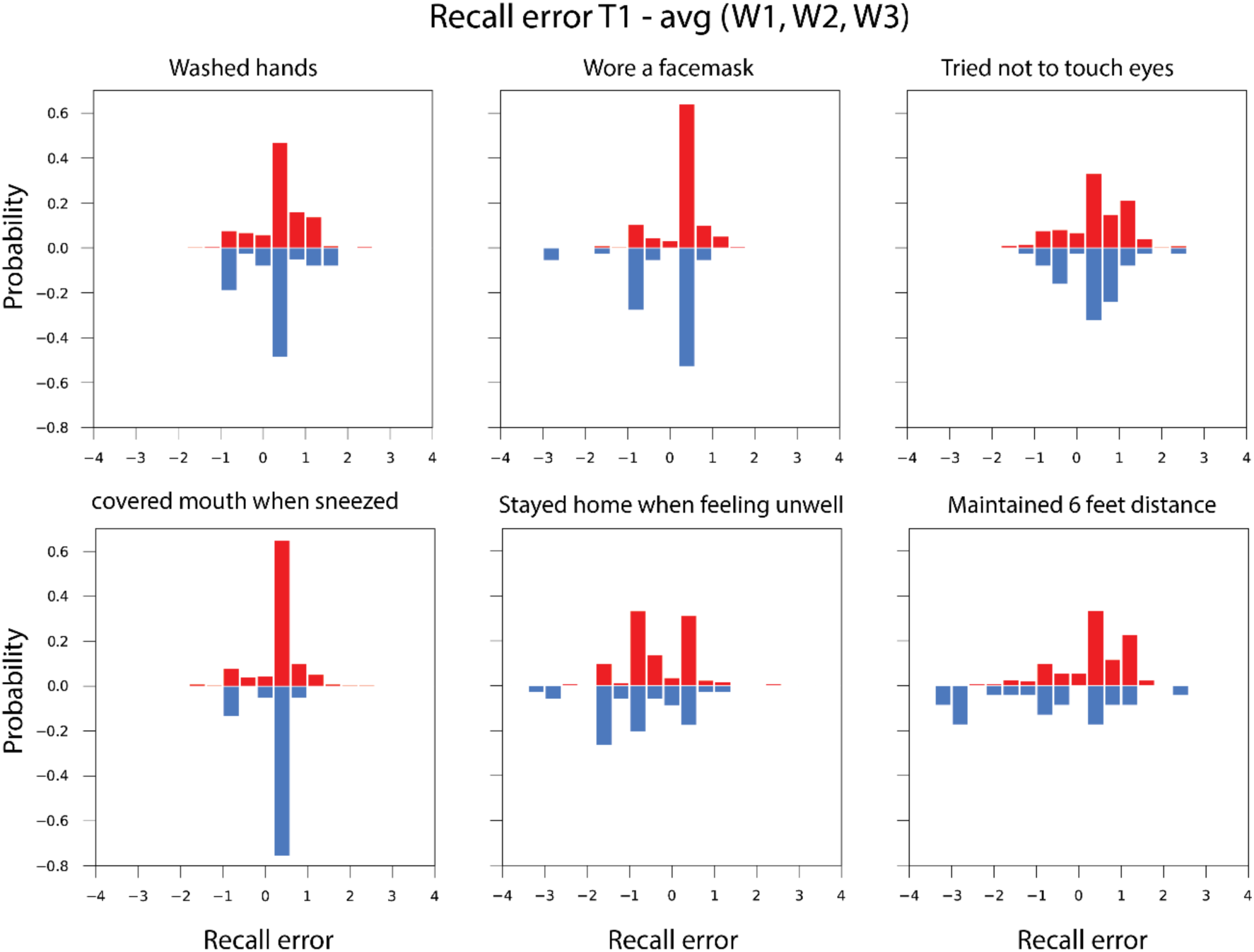
The distribution of differences in individual behaviors for students (blue) and non-students (right) between T1 based on W5 recall and the weighted average of individual behaviors collected in real time during W1-W3. The skewness of the distribution influences whether a previous behavior is understated (skewed to the left) or overstated (skewed to the right). The analysis reveals that students more often understated their previous protective behaviors, while non-students more often overstated their previous protective behaviors.

Among these six behaviors, we anticipate no significant impact on results for wearing facemasks, washing hands, avoiding touching eyes, and covering the mouth when sneezing as well as maintaining six feet of distance from others among non-students. This is because, for those behaviors with significant recall bias, the average recall error never exceeded ±0.22 units on a scale of 0 to 4 and did not result in cluster membership change. For maintaining six feet among students, the average recall error was −0.98, indicating that students understated their compliance with this behavior in T1 during surveys in W5. Recall bias for staying home when feeling unwell could have significant implications for results. This behavior was understated by both cohorts in T1, with an average recall error of −1.14 for students and −0.58 for non-students. The implication is that both cohorts stayed home when sick during T1 more than in subsequent time frames, resulting in a slightly declining behavior trajectory. This suggests a potential but modest role for this behavior in disrupting the 2020-2021 influenza season. For the student cohort, we advise caution when interpreting average differences, as they are based on a small population size (n=37) consisting of students in W1, W2, W3, W4, and W5.

